# Human Epidemiology and RespOnse to SARS-CoV-2 (HEROS): Objectives, Design and Enrollment Results of a 12-City Remote Observational Surveillance Study of Households with Children using Direct-to-Participant Methods

**DOI:** 10.1101/2022.07.09.22277457

**Authors:** Patricia C. Fulkerson, Stephanie J. Lussier, Casper G. Bendixsen, Sharon M. Castina, Tebeb Gebretsadik, Jessica S. Marlin, Patty B. Russell, Max A. Seibold, Jamie L. Everman, Camille M. Moore, Brittney M. Snyder, Kathy Thompson, George S. Tregoning, Stephanie Wellford, Samuel J. Arbes, Leonard B. Bacharier, Agustin Calatroni, Carlos A. Camargo, William D. Dupont, Glenn T. Furuta, Rebecca S. Gruchalla, Ruchi S. Gupta, Gurjit Khurana Hershey, Daniel J. Jackson, Christine C. Johnson, Meyer Kattan, Andrew H. Liu, Liza Murrison, George T. O’Connor, Wanda Phipatanakul, Katherine Rivera-Spoljaric, Marc E. Rothenberg, Christine M. Seroogy, Stephen J. Teach, Edward M. Zoratti, Alkis Togias, Tina V. Hartert, the HEROS study team

**Author notes:** Corresponding author: Patricia C. Fulkerson, MD, PhD, National Institutes of Allergy and Infectious Diseases, 5601 Fishers Lane, 6B56, Bethesda, MD 20892 MSC9827. DISCLAIMER: Drs. Fulkerson’s and Togias’ authorship does not constitute endorsement by the National Institute of Allergy and Infection Diseases, the National Institutes of Health or any other Agency of the United States Government. **Meeting presentation** Study methods and findings were presented at the American Thoracic Society meeting, San Francisco, CA, USA, May 2022.

## Abstract

The Human Epidemiology and Response to SARS-CoV-2 (HEROS) is a prospective multi-city 6-month incidence study which was conducted from May 2020-February 2021. The objectives were to identify risk factors for SARS-CoV-2 infection and household transmission among children and people with asthma and allergic diseases, and to use the host nasal transcriptome sampled longitudinally to understand infection risk and sequelae at the molecular level. To overcome challenges of clinical study implementation due to the coronavirus pandemic, this surveillance study used direct-to-participant methods to remotely enroll and prospectively follow eligible children who are participants in other NIH-funded pediatric research studies and their household members. Households participated in weekly surveys and biweekly nasal sampling regardless of symptoms. The aim of this report is to widely share the methods and study instruments and to describe the rationale, design, execution, logistics and characteristics of a large, observational, household-based, remote cohort study of SARS-CoV-2 infection and transmission in households with children. The study enrolled a total of 5,598 individuals, including 1,913 principal participants (children), 1,913 primary caregivers, 729 secondary caregivers and 1,043 other household children. This study was successfully implemented without necessitating any in-person research visits and provides an approach for rapid execution of clinical research.

## Introduction

The Human Epidemiology and Response to SARS-CoV-2 (HEROS) study is a prospective 12-city 6-month incidence study of SARS-CoV-2 within households with children. The study was initiated in May 2020, under national lockdown conditions, which rendered even surveillance studies exceedingly difficult to conduct. To overcome these limitations HEROS employed direct-to-participant methods to remotely enroll and prospectively follow US households across the nation, including the self-collection of child and adult biosamples.

This study was motivated by the recognition that at the start of the SARS-CoV-2 pandemic, by necessity, studies were limited to individuals who were tested for SARS-CoV-2 and those with severe disease. However, the role of children was not a focus at the beginning of the pandemic as they were generally spared from severe disease. As a result, information on the rate at which children were infected and their potential role in silent transmission was unknown (1). At the start of the pandemic, asthma was not identified as a risk factor for severe infection, which was unexpected, as people with asthma typically experience significant morbidity from many respiratory viruses and are target groups for vaccine preventable respiratory viral diseases (2). Additionally, the role of other allergic diseases as risk factors was unknown.

To address these gaps, the National Institute of Allergy and Infectious Diseases (NIAID) assembled a consortium of experienced clinical sites and, in collaboration with their investigators, designed and implemented the HEROS surveillance study. In addition to obtaining more information on the role of children in the pandemic, HEROS aimed at determining whether people with asthma and allergic diseases are at increased risk for SARS-CoV-2 infection and evaluating whether the host nasal transcriptome longitudinally associated with infection and transmission risk. The aim of this manuscript is to share these methods and to describe the rationale, design, execution, logistics and characteristics of a large, observational, household-based, remote cohort study of SARS-CoV-2 infection and transmission in households with children across the United States.

## Methods

### Study objectives

HEROS was designed as a prospective, observational study in which children that were currently enrolled or had been enrolled in National Institute of Health (NIH)-funded asthma and allergic disease-focused studies and their household contacts were surveyed with weekly questionnaires and biweekly nasal collections over the study period from May 2020 to February 2021 during a period prior to vaccine availability. The primary objective was to determine the incidence of SARS-CoV-2 infection via detection of viral RNA in nasal secretions. Details on HEROS study objectives and methods are included in supplementary materials. The study was registered at ClinTrials.gov where more details of the study protocol are described [NCT04375761].

### Public health surveillance exception

The Vanderbilt University Institutional Review Board (IRB), which was designated as the study single IRB, reviewed the study protocol and determined that the study satisfied criteria for the public health surveillance exception [45CFR46.102(I)(2)]. In assessing this determination, NIAID staff conferred with the Department of Health and Human Services Office for Human Research Protections (OHRP) regarding the applicability of the public health surveillance exception for the HEROS study protocol. OHRP concurred with the public health surveillance determination by the Vanderbilt IRB for the HEROS protocol. The Vanderbilt IRB shared the determination with all participating sites in reliance and all participating investigators were required to confirm the public health surveillance exception for the HEROS protocol from their respective institutional IRB before site activation. The primary household caregiver was provided an online study information fact sheet, a consent-like document containing components of the common rule. The primary caregivers were required to agree that they read, understood, and reviewed this information sheet with all participating household members in order to enroll, including an option to select having the study site contact them to answer any questions.

### Participants and enrollment

The cohorts from which HEROS households were recruited included population-based cohorts, as well as disease-specific cohorts (Supplemental Table 1). An eligible household had to include a 21-years-old and younger individual who was or had been a participant in a NIH-funded asthma and other allergic diseases study (principal participant) and who was expected to remain in the United States with a caregiver for the duration of the study. A minimum of 2 participants (principal participant and a caregiver) and a maximum of 4 participants (a second caregiver and/or a sibling under 21 years of age residing in the same household) per household were eligible to enroll. To be eligible, the principal participant needed to live with the caregiver for at least 50% of the time during the study period. Principal participants and their household contacts were recruited from 20 established NIH-funded cohorts in 12 cities, including 13 individual clinical sites, across the U.S. (Figure 1).

**Figure 1.**
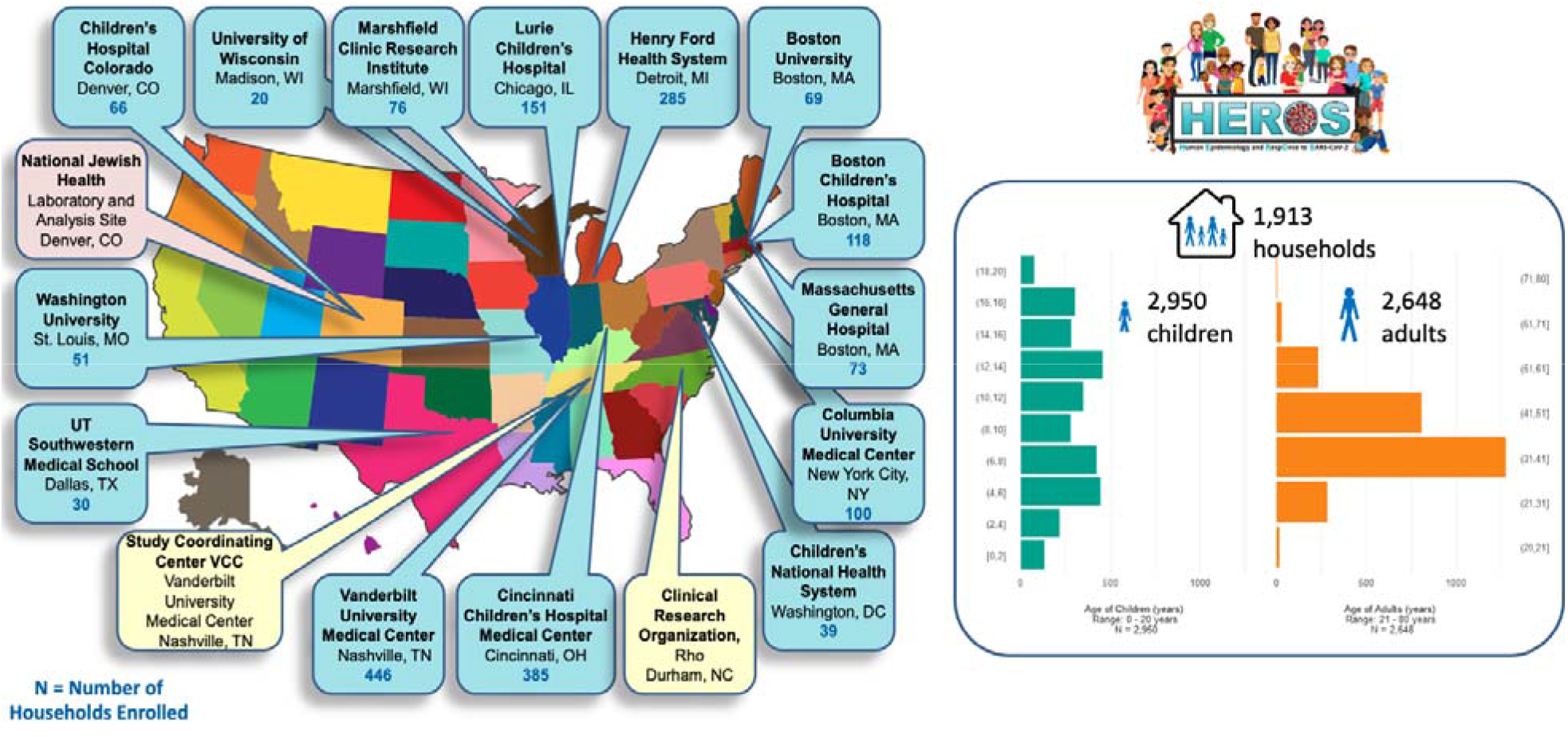
HEROS clinical, laboratory and coordinating sites. The left panel depicts the clinical, laboratory and coordinating study sites. The right panel summarizes the enrolled households, numbers of adults and children, and their age distribution.

The goal was to enroll 2000 households over approximately 8 weeks. Participants were recruited through emails, letters, and phone calls from the HEROS clinical sites. After review and agreement with the study fact sheet, a caregiver completed a baseline registration form and an enrollment questionnaire, which collected demographic, medical history, and current medication information for each participating member of the household and information on the household environment. A majority (94%) of the baseline enrollment questionnaires were completed on-line by the caregiver, yet study staff did assist a small fraction (6%) of the caregivers with baseline questionnaire completion over telephone communication. As part of the registration form, the primary contact person (caregiver 1 [CG1]) was identified for the household. Designed like the United States census, with which families would have recently completed and would likely be familiar, CG1 completed the surveys on behalf of all participating household members (3). A household was considered enrolled in the study after completion of the registration form and the baseline questionnaire and all participating household members were then assigned unique study participant numbers. Among enrolled households, a household was considered completed if the exit survey was completed. If no exit survey was completed, the household was considered withdrawn (or terminated) from the study. Notably, data from all participants, including participants from households that did not fully complete the study, are included in analyses unless otherwise stated (4). Each enrolled household participated in the study for 24 weeks with an option to participate for an additional 4 weeks (maximum 28 weeks). The extension was added, as the study was being conducted in real-time during the pandemic, and the extension included coverage of what was the fall and winter COVID surges during 2020.

### Data collection instruments

All study materials were provided in English and Spanish, and study participants could select their preferred language. Surveys to capture symptoms and illnesses were completed weekly and surveys to collect household exposure information were completed every other week (Figure 2). All surveys were completed remotely via a smart phone, on-line, or a telephone communication with local study site staff.

**Figure 2.**
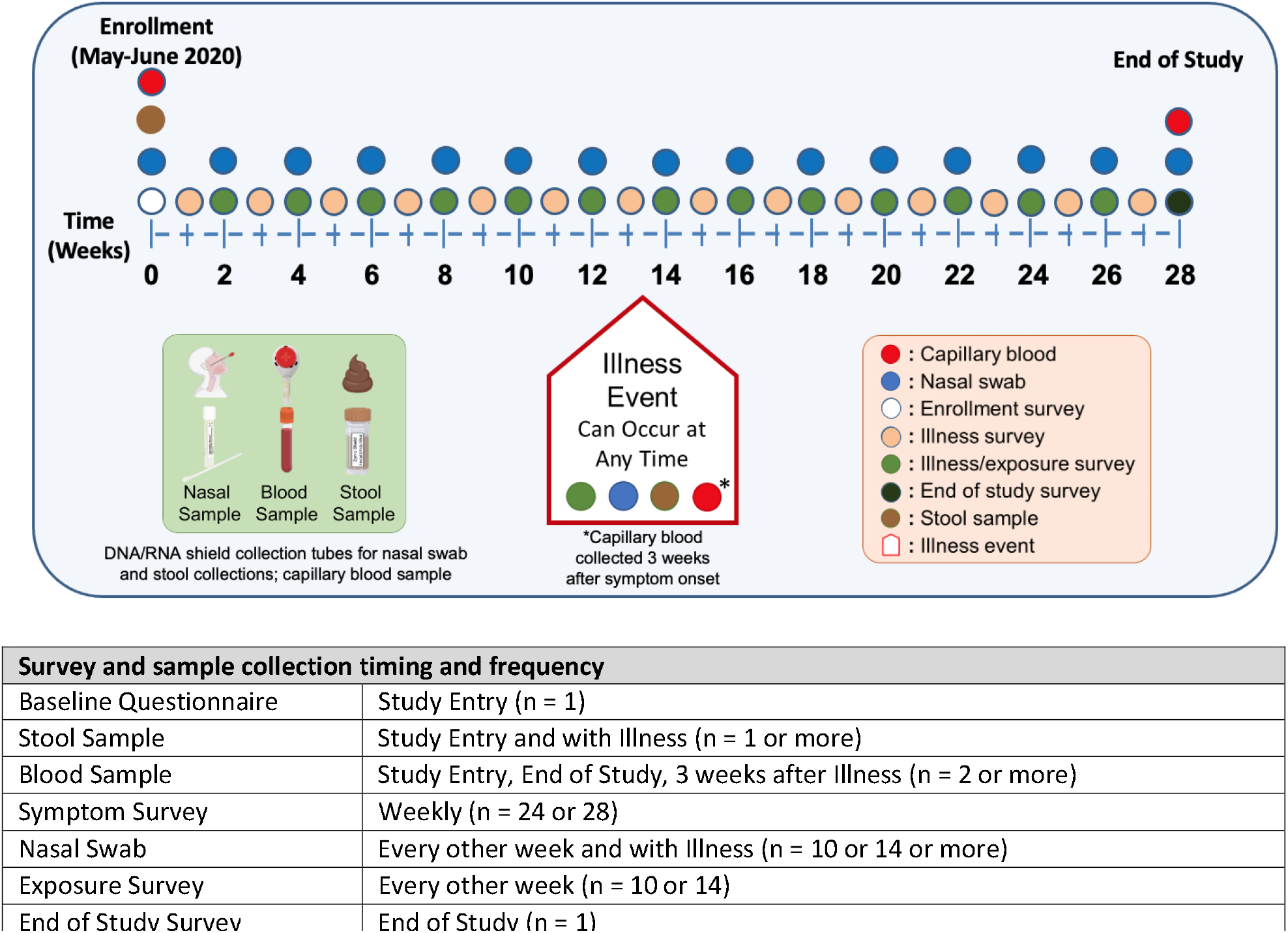
HEROS study survey and sample collection timetable and study enrollment of households, adults and children. The schematic of the HEROS study design depicts the timeline of events with enrollment data and biospecimen collection, weekly surveys, biweekly nasal swabs, and additional illness visits triggered by an automated algorithm based on clinical symptom survey responses.

For those households that indicated a preference for communication via text or email, a link to the electronic surveys was sent to CG1 every week. If an illness in the household was reported on the weekly survey, CG1 completed a symptom survey for the affected household member. A symptom algorithm was designed and integrated into the data capture to identify participants with symptoms compatible with SARS-CoV-2 infection (Supplemental Figure 1). If a study-specified symptom threshold was met (Supplemental Figure 1), an illness event was triggered for the household and CG1 was instructed to collect additional illness-associated samples from all enrolled household participants.

The HEROS baseline questionnaire and surveys were primarily developed using items from previously validated or widely used survey instruments (5-13). The enrollment survey collected information on basic demographics and household features, and the enrollment and biweekly surveys included assessment of factors associated with exposure risks and risk-taking behaviors. A list of exposures was asked at the household level at enrollment and biweekly. The HEROS survey instruments are available at https://www.vumc.org/heros/survey-instruments and the online supplement.

Data were entered into a database via a study-specific electronic data capture system (https://project-redcap.org, Vanderbilt University, Nashville, TN) and then transferred daily to the Division of Allergy, Immunology and Transplantation Statistical and Clinical Coordinating Center (DAIT SACCC, Rho Inc., Durham, NC) by an application programming interface (API). Raw data from the REDCap database was transformed into analyses datasets which were used for all analyses.

### Remote biospecimen collection

Supplemental Figure 2 is a schematic of the data and biospecimen collection and management. After completion of the baseline questionnaire, an initial sample collection kit was shipped overnight to the participating households. This contained written instructions and materials needed for home collection of biological specimens, including nasal swab, capillary blood, and stool. Participants were instructed to watch training videos before performing nasal swabs and blood collection at home, and as needed, study site personnel assisted families with collections via video conferencing or telephone calls. Capillary blood collection from participants ages 2 and older was done at home using provided capillary blood collection devices for serum separation (Tasso, Inc, Seattle, WA). The home collection kits also contained personal protective equipment, including masks and gloves, and mailers for return postal shipment at ambient temperature of the collected biospecimens to a central repository.

Nasal swabs were collected from all household members that were enrolled in the study starting in the second week and every 2 weeks thereafter for a total of 12 swabs collected over 24 weeks (Figure 2). If the household agreed to participate in the study extension, additional nasal swabs were collected for a total of 28 weeks. Swab collection tubes were prefilled with a reagent that stabilized the sample and neutralized any virus (Zymo Research Corporation, Irvine, CA). For each collection, sufficient sterile nasal swabs and pre-labeled swab collection tubes were provided together in a bag. Baseline and illness-associated stool samples were collected via swabs of stool on toilet paper and placed into the same type of prefilled swab collection tubes as used for nasal sampling but marked for stool. After collection, the nasal and stool swabs were broken off into the prefilled collection tubes, which were then placed into absorbent pouches and then into biohazard bags and box mailers for shipment to the study biorepository. Capillary blood samples were collected from the upper arm or lower back from participating household members 2 years of age and older at study start, 3 weeks after an illness event, and at the end of study. Details on sample collection and tracking are included in Supplemental Methods.

### Defining SARS-CoV-2 infection

PCR testing for SARS-CoV-2 was conducted on nasal and stool swabs using the Centers for Disease Control (CDC) SARS-CoV-2 N1, N2, and RNaseP housekeeping gene assays. Stool samples were tested using modified RNA purification and thermocycling protocols optimized for detection of SARS-COV-2 RNA in stool (DOI: 10.1016/S2666-5247(21)00059-8). The assays using nasal swabs were run in duplicate and SARS-CoV-2 positivity was defined as having at least 2 of the 4 N1 and N2 assays amplify with a Cq < 40, through any combination of assays. Viral Cq values were normalized to RNase P expression levels for each assay N1 and N2 and transformed from log_2_ scale into viral load values (viral load(N_x_) = 2^Cq(RNaseP) – Cq(Nx)^ where N_x_ is either N1 or N2), then averaged across N1 and N2 assays to generate a relative viral load value for each sample.

### Defining asthma and allergic diseases

Asthma, allergic rhinitis, atopic dermatitis and food allergy were defined by self-report of physician diagnosis. Medical and medication history were assessed at enrollment and end of study. In order to assess the level of IgE sensitization to common aeroallergens and foods, the ImmunoCAP ISAC method (Portage, MI), a multiplex specific IgE test for 112 allergen components from 48 different allergen sources, was used on serum from capillary blood (14).

### Study component completion and cohort retention

In addition to automated reminders to complete study components, study sites received weekly reports of their site’s HEROS participants to allow study staff to directly contact participating households who were not completing surveys and/or biospecimen collection. The families were compensated for their time, sent HEROS materials that were deemed useful during the pandemic to encourage engagement (masks, playing cards, pins and stickers), and sent a monthly HEROS newsletter to update them on the study progress and results as they became available.

## Results

The study was designed and implemented in 6 weeks from the start of protocol development to the enrollment of the first household on May 1, 2020. From May 2020 through June 2020, among 6,160 eligible households currently active in participating cohorts whose study consent allowed contact for other studies, 1,963 households were registered into the study with 1,913 households in 11 states completing the baseline questionnaire for enrollment (Figure 3). The study enrolled a total of 5,598 individual participants, which included: 1,913 principal participants, 1,913 primary caregivers (CG1), 729 secondary household adults or caregivers and 1,043 siblings or other household children. The average size of the participating households was 4.4 individuals, and the average number or participants enrolled in HEROS per household was 2.9. The primary caregiver was the mother (94%), father (4%), and grandmother or other household member (2%). The mean age of the principal participant was 10.5 years (s.d. 4.8). The age distribution is shown in Figure 1. The principal participant race was white (54%), Black or African American (31%), more than one race (6%), preferred not to answer (7%), or other race (2%).

**Figure 3.**
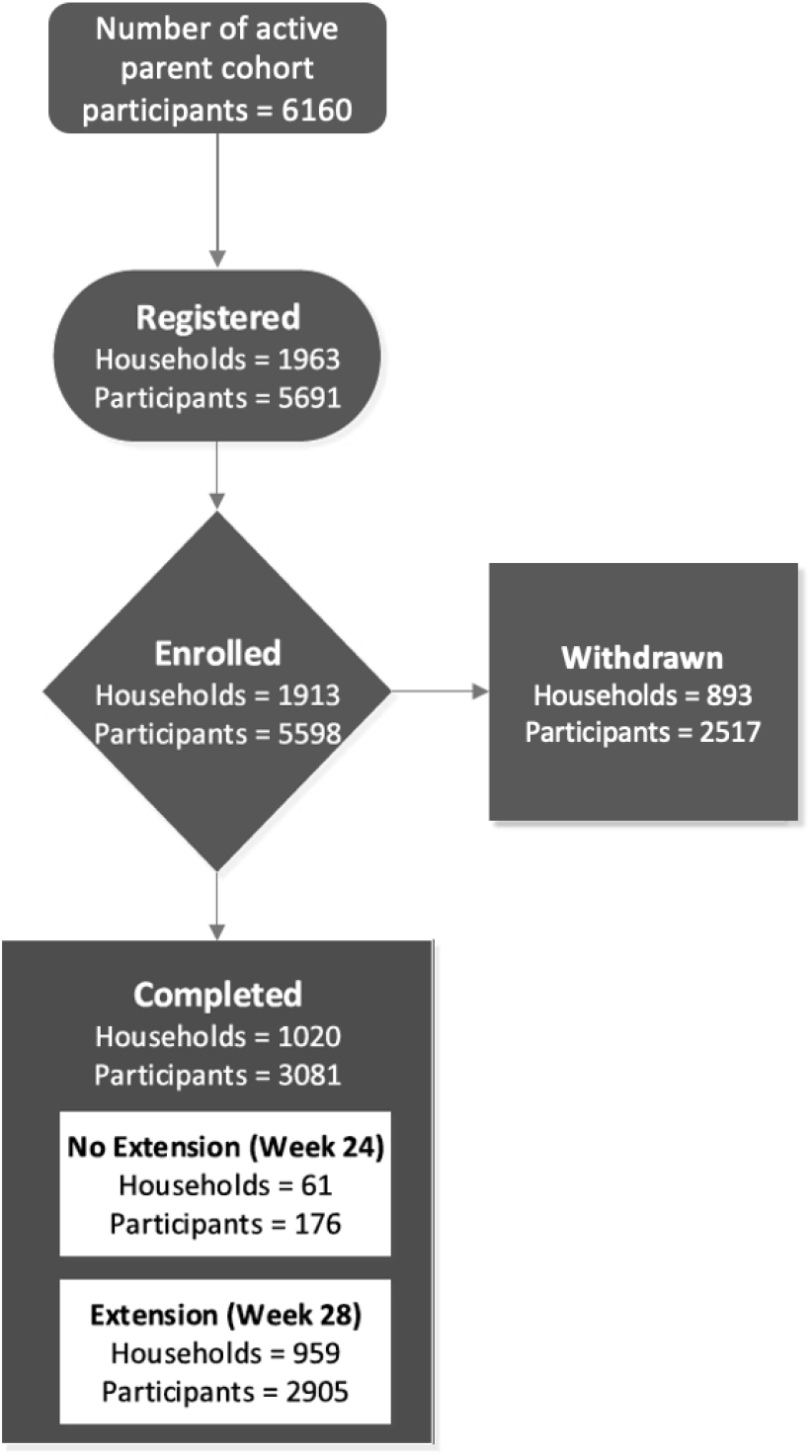
HEROS study flow diagram of parent cohort participants, and household and individual participant enrollment through 6-month study completion.

Among the originally enrolled 1,913 households, 893 households were withdrawn from the study (Table 1). Eighty percent of these households (712/893) were withdrawn due to non-completion of surveys and sample collection following enrollment. Another major reason for withdrawal from the study was participant elected to withdrawal (150/893, 18%). A comparison of the demographics of completed versus withdrawn households is shown in Table 1, and Tables 2 and 3 provide individual level demographics of all participants and caregiver participants, respectively. Self-reported household and clinical and social characteristics are presented in Supplemental Table 2. Notably, a larger proportion of black or African American participants enrolled in HEROS were withdrawn as compared to white participants (Table 2). In addition, age and race of the caregivers were significant when we compared caregivers from households that were withdrawn compared to those that completed the study (Table 3).

**Table 1.** Household characteristics.

| **Table 1. Household characteristics** |  |  |  |
| --- | --- | --- | --- |
|  | **Completed,**<br>**N = 1,020<sup>1</sup>** | **Withdrawn,**<br>**N = 893<sup>2</sup>** | **Overall,**<br>**N = 1,913<sup>1</sup>** |
| **Number of Enrolled Household Members** |  |  |  |
| 2 | 362 (35%) | 378 (42%) | 740 (39%) |
| 3 | 275 (27%) | 299 (33%) | 574 (30%) |
| 4 | 383 (38%) | 216 (24%) | 599 (31%) |
| **Household Size** | 4.38 (1.31) | 4.35 (1.51) | 4.37 (1.41) |
| **Number of Bedrooms** | 3.47 (1.06) | 3.31 (1.05) | 3.39 (1.06) |
| **Members / Bedrooms** | 1.36 (0.56) | 1.39 (0.53) | 1.37 (0.55) |
| N Missing | 1 | 2 | 3 |
| **Pets in Home** | 588 (58%) | 472 (53%) | 1,060 (55%) |
| **Preferred Method of Contact** |  |  |  |
| Email | 353 (35%) | 226 (25%) | 579 (30%) |
| Phone Call | 53 (5.2%) | 91 (10%) | 144 (7.5%) |
| Text Message | 613 (60%) | 574 (64%) | 1,187 (62%) |
| N Missing | 1 | 2 | 3 |
| **Preferred Language** |  |  |  |
| English | 992 (97%) | 868 (97%) | 1,860 (97%) |
| Spanish | 27 (2.6%) | 23 (2.6%) | 50 (2.6%) |
| N Missing | 1 | 2 | 3 |
| **Reported Any Household Hospitalized for COVID-19** | 4 (0.4%) | 8 (0.9%) | 12 (0.6%) |
| **Reported high-risk COVID-19 condition** | 669 (66%) | 647 (72%) | 1,316 (69%) |
| <sup>1</sup>n (%); Mean (SD)<br><sup>2</sup>High risk medical conditions include: asthma, autoimmune conditions, cancer, COPD, congestive heart failure, coronary artery or coronary heart disease, cystic fibrosis, emphysema, heart attack, hypertension or high blood pressure, type I diabetes, or type II diabetes. |  |  |  |

**Table 2.** Individual participant characteristics.

|  | Completed,<br>N = 3,081<sup>1</sup> | Withdrawn,<br>N = 2,517<sup>2</sup> | Overall,<br>N = 5,598<sup>2</sup> | p-value<sup>2</sup> |
| --- | --- | --- | --- | --- |
| **Age (Participants < 20 Years)** | 10.0 (6.0, 14.0) | 11.0 (7.0, 14.0) | 10.0 (6.0, 14.0) | <0.001 |
| **Age (Participants 21 and Older)** | 40 (36, 46) | 39 (34, 45) | 40 (35, 45) | <0.001 |
| **Sex (Participants Less Than 20 Years)** |  |  |  | 0.561 |
| Male | 834 (52%) | 698 (52%) | 1,532 (52%) |  |
| Female | 756 (47%) | 651 (48%) | 1,407 (48%) |  |
| Other | 7 (0.4%) | 3 (0.2%) | 10 (0.3%) |  |
| **Sex (Participants 21 and Older)** |  |  |  | <0.001 |
| Female | 1,011 (68%) | 899 (77%) | 1,910 (72%) |  |
| Male | 468 (32%) | 264 (23%) | 732 (28%) |  |
| Other | 4 (0.3%) | 0 (0%) | 4 (0.2%) |  |
| **Race** |  |  |  | <0.001 |
| White or Caucasian | 2,137 (69%) | 1,100 (44%) | 3,237 (58%) |  |
| Black or African American | 518 (17%) | 1,071 (43%) | 1,589 (28%) |  |
| Prefer Not to Answer | 197 (6.4%) | 199 (7.9%) | 396 (7.1%) |  |
| More than One Race | 128 (4.2%) | 101 (4.0%) | 229 (4.1%) |  |
| Asian | 69 (2.2%) | 21 (0.8%) | 90 (1.6%) |  |
| American Indian or Alaska Native | 26 (0.8%) | 20 (0.8%) | 46 (0.8%) |  |
| Other | 6 (0.2%) | 5 (0.2%) | 11 (0.2%) |  |
| **Ethnicity** |  |  |  | 0.038 |
| Non-Hispanic | 2,669 (87%) | 2,132 (85%) | 4,801 (86%) |  |
| Hispanic | 353 (11%) | 312 (12%) | 665 (12%) |  |
| Prefer Not to Answer | 58 (1.9%) | 71 (2.8%) | 129 (2.3%) |  |
| Other | 1 (<0.1%) | 2 (<0.1%) | 3 (<0.1%) |  |

<sup>1</sup> Median (IQR); n (%)<sup>2</sup> Wilcoxon rank sum test; Fisher's Exact Test for Count Data with simulated p-value (based on 2000 replicates)

**Table 3.** Caregiver characteristics.

|  | Completed, N<br>= 1,020<sup>1</sup> | Withdrawn,<br>N = 893<sup>1</sup> | Overall,<br>N = 1,913<sup>1</sup> | p-value<sup>2</sup> |
| --- | --- | --- | --- | --- |
| **Age** | 40 (36, 46) | 38 (34, 44) | 39 (35, 45) | <0.001 |
| **Sex** |  |  |  | 0.402 |
| Female | 962 (94%) | 855 (96%) | 1,817 (95%) |  |
| Male | 56 (5.5%) | 37 (4.1%) | 93 (4.9%) |  |
| Other | 2 (0.2%) | 1 (0.1%) | 3 (0.2%) |  |
| **Race** |  |  |  | <0.001 |
| White or<br>Caucasian | 708 (69%) | 385 (43%) | 1,093 (57%) |  |
| Black or African<br>American | 186 (18%) | 394 (44%) | 580 (30%) |  |
| Prefer Not to<br>Answer | 71 (7.0%) | 73 (8.2%) | 144 (7.5%) |  |
| More than One<br>Race | 21 (2.1%) | 24 (2.7%) | 45 (2.4%) |  |
| Asian | 26 (2.5%) | 7 (0.8%) | 33 (1.7%) |  |
| American<br>Indian or Alaska<br>Native | 7 (0.7%) | 8 (0.9%) | 15 (0.8%) |  |
| Other | 1 (<0.1%) | 2 (0.2%) | 3 (0.2%) |  |
| **Ethnicity** |  |  |  | 0.186 |
| Non-Hispanic | 888 (87%) | 759 (85%) | 1,647 (86%) |  |
| Hispanic | 114 (11%) | 107 (12%) | 221 (12%) |  |
| Prefer Not to<br>Answer | 18 (1.8%) | 26 (2.9%) | 44 (2.3%) |  |
| Other | 0 (0%) | 1 (0.1%) | 1 (<0.1%) |  |

<sup>1</sup>Median (IQR); n (%)<sup>2</sup>Wilcoxon rank sum test; Fisher's exact test for count data with simulated p-value (based on 2000 replicates)**Biospecimen sample collection**

### Surveys

Links to surveys were offered via multiple modalities; 60% chose texting for surveys, 35% email, and 5% phone. Among the 1,020 households that completed the study, 28,011 weekly surveys were distributed with 90% response. There were 14,158 biweekly surveys distributed and 92% were completed. Among all the enrolled households, 1451 illness events were identified from 904 study participants with 61% of the illness nasal samples collected.

### Biospecimen sample collection

Among the 1020 households that completed the HEROS study, biweekly nasal swabs were received by the sample repository for 76% of scheduled collections. Supplemental Table 3 shows the biospecimen collection adherence by caregiver demographics, and the nasal sample collection over study time is shown in Supplemental Figure 3. The quality of the nasal samples was assessed by nasal swab qPCR of the RNaseP housekeeping gene, which was successful in 99.9% of samples. Baseline capillary blood was collected from 82% (N=3030) of eligible (age 2+ years) participants at enrollment, and stool from 85% of eligible participants. End of study capillary blood was collected from 66% (N=3030) of eligible (age 2+ years) participants. There was sufficient serum volume from returned capillary collection devices for at least one aliquot of 30 mcL in 81% of returned collection tubes.

### Participant survey on HEROS study experience

The end of study survey included questions regarding the HEROS study and remote biospecimen collections. A summary of the participant experience and responses is in Table 4. The video demonstrating sample collection was recognized as helpful for 94% of the households that completed the study and as most helpful for successful sample collection for more than half (Table 4). In addition, 89% of the households that completed the study felt confident using the capillary blood collection device at home after reviewing the instructions and/or videos and, notably, 89% of the households would choose to use the device for blood collection in future studies.

**Table 4.**
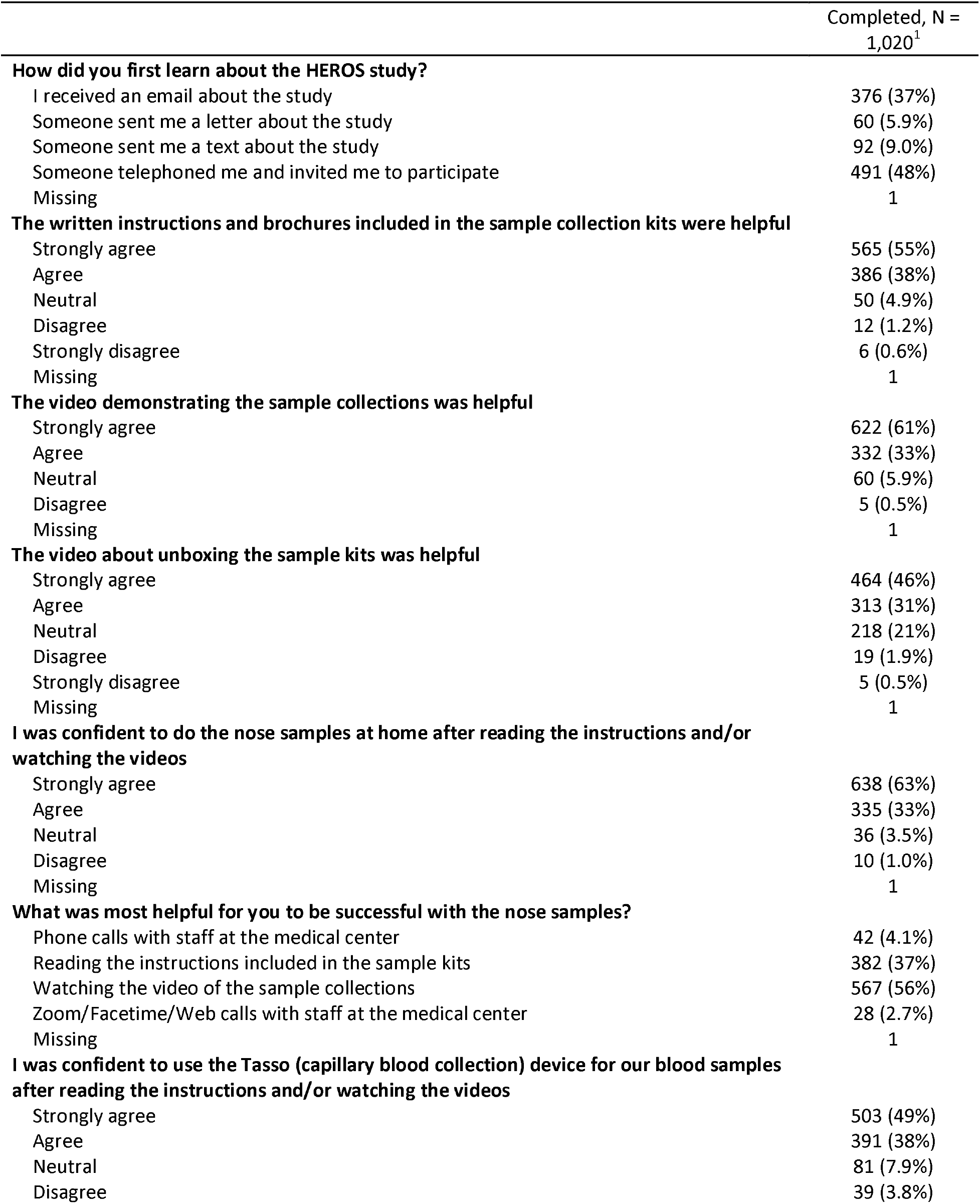
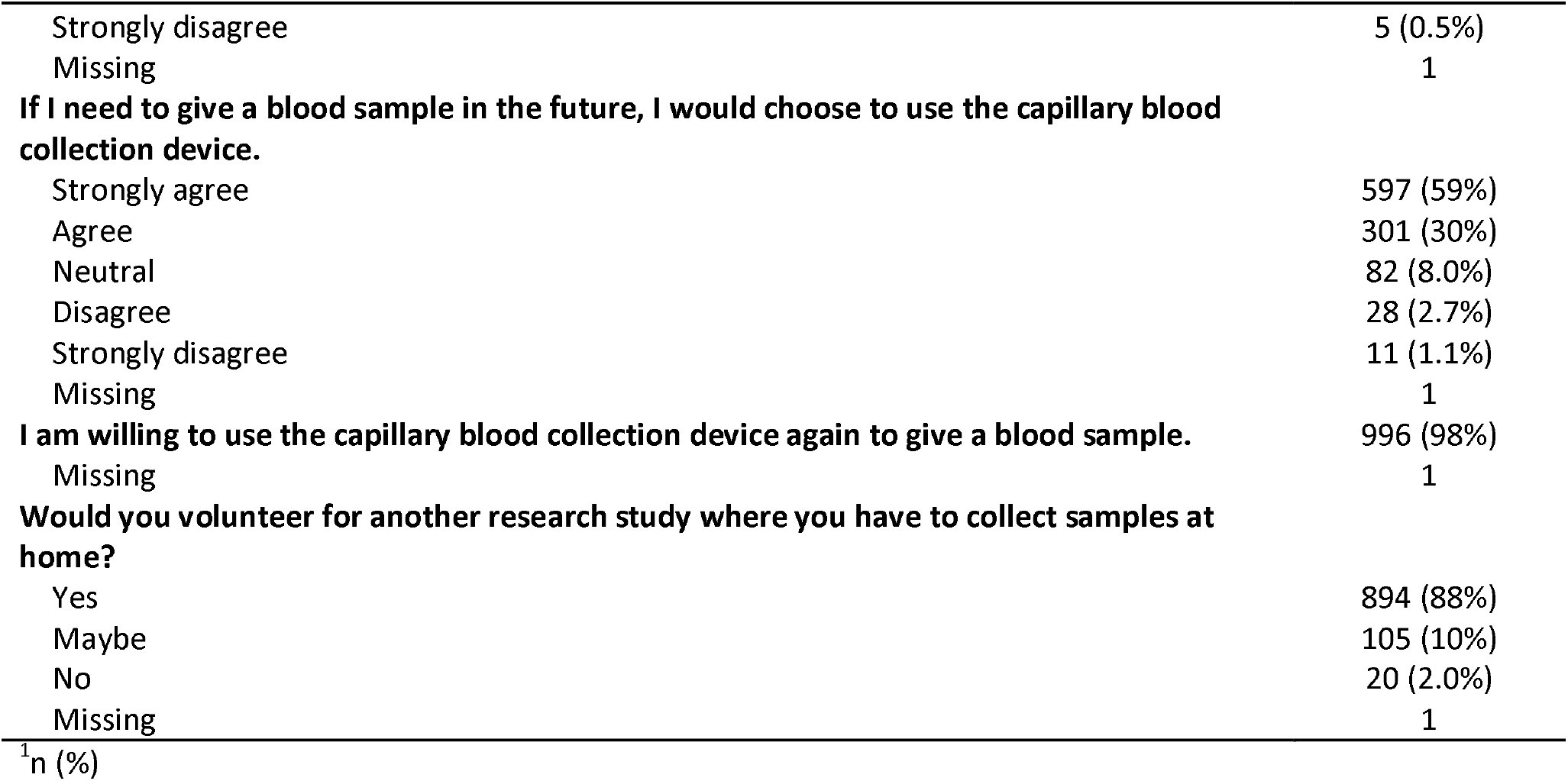
Household study experience assessed at end of study.

### Adverse events

Adverse events (AEs) were recorded and were uncommon (N=25 in 19 participants). Most adverse events occurred in the principal child (72%). The most common AEs were related to the capillary blood collection and included irritation from the adhesive and bruising at the adhesive site. Of the 25 AEs reported, there were 3 Grade 3 events in one participant that experienced syncope and a fall causing injuries, including a concussion and fracture of facial bones, during a capillary blood collection. Supplemental Table 4 provides a summary of the adverse events.

## Discussion

The HEROS study was designed to address important gaps in our understanding of the role of children and those with asthma and allergic diseases in SARS-CoV-2 infection risk and transmission. HEROS enrolled households with children who were participating or had previously participated in 20 NIH cohorts focused on asthma and allergic diseases in 12 cities.

The need for a rapid transition to direct-to-participant research studies was akin to the rapid transition to telehealth during the pandemic. The benefit of recruiting families enrolled in other studies was that they would be familiar with research methods and nasal sampling, allowing rapid roll-out of an entirely direct-to-participant study. This was a novelty and HEROS’ successful recruitment proved the value of this approach. In addition, the following elements were probably important in the success of HEROS: a) study participants could select how they would complete study surveys, with most selecting to complete them electronically, most commonly using a smartphone; b) the training on collection of the biospecimens was multi-faceted and included written materials that accompanied the kits, videos, and contact with the study site research team for video or phone conferencing. Overall, from the HEROS caregivers’ responses to the exit questionnaire, it is clear that the concept of at-home biospecimen collection in clinical trials can be substantially extended to include respiratory (nasal) sampling as well as capillary blood collection in studies where these specimens are of value.

The most important obstacle we encountered in HEROS was the high drop-out rate. While families were willing to participate and a large number was originally enrolled, it quickly became apparent that active participation would end up being smaller. Specific problems that we believe culminated in this reduction included a) significant delays in distributing the biosample kits due to lock-down associated difficulties with availability of materials such as PPEs, swabs, reagent tubes, kit assembling and shipping, b) relatively complicated instructions to use the contents of the sample kits, c) some inconsistency with clinical site staff contacting participants and assisting with the completion of questionnaires and biosample collection and d) perhaps most importantly, the overall demands of the study requiring repetitive sampling and survey completion in the middle of an unprecedented health and social crisis. Whether with additional assistance, or even home visits, more households could have completed the study is not known, but the characteristics of those who remained in the study compared to those who exited, suggest that additional retention methods are needed to engage families in direct-to-participant remote studies, particularly for longitudinal studies requiring frequent surveys and biospecimen collections. Another obstacle that HEROS encountered was a prolonged shipping time for biospecimens to arrive at the central repository from participating households. The impact of this delay on nasal swabs seems to have been minimal judging from the ability to amplify RNA from 99.9% of samples, but the quality of blood specimens was affected in that hemolysis was common. Fortunately, the serologic assays that were ultimately performed were not influenced by this problem.

HEROS focused on a convenience sample in order to enrich the study with participants who had asthma and other allergic diseases in existing populations of ongoing or previously completed NIH studies. The determination of doctor-diagnosed asthma and allergic conditions was made through self-report, which can result in misclassification. To overcome this, asthma and allergic disease medications were collected both at enrollment and at the end of study, and blood specimens were used to measure specific IgE to aeroallergens and food. As the study built upon existing cohorts focused on asthma and allergic diseases, these were households who were familiar with research, and many with nasal collections, although none with entirely remote studies. Therefore, whether participation would be generalizable to other groups, particularly those unfamiliar with research, is unknown. In addition, there was a strong sense of purpose among study families at the beginning of the pandemic, and this sense of engagement may not translate to remote research methods in other times and settings. Another limitation derives from the 14-day time window chosen for the nasal biospecimen collections. Although this choice was based on known viral shedding duration balanced with our effort to avoid excessive study fatigue, asymptomatic infections that began and resolved between regular biospecimen collections could theoretically have been missed. Also, because of the lack of more dense sampling, it was not possible to identify the index case within a household with multiple concurrent infections.

In summary, the HEROS study successfully employed a direct-to-participant research strategy which was necessary given that in-person research visits could not be done during the early pandemic, precluding direct involvement of research staff. We demonstrated the ability to rapidly implement a remote methods study including both survey as well as nasal, capillary blood and stool biospecimen collections in 12 cities across the US. The successes and obstacles encountered, and the overall experience obtained from HEROS will be useful not only for deployment of similar studies in the future, but also for improving recruitment and retention of participants in conventional studies, where reduction in the number of on-site visits with at-home biospecimen collection can be of substantial value.

## Data Availability

Requests may be made for use of data from the HEROS study to the NIAID. A protocol committee will review requests as use of HEROS data is restricted to studies focused on SARS-CoV-2 as this study was conducted as public health surveillance of SARS-CoV-2.

## Abbreviations

IQR: (interquartile range)
PCR: (polymerase chain reaction)
SARS-CoV-2: (severe acute respiratory syndrome coronavirus 2)
SD: (standard deviation)

## Acknowledgements

**This work was supported by:**

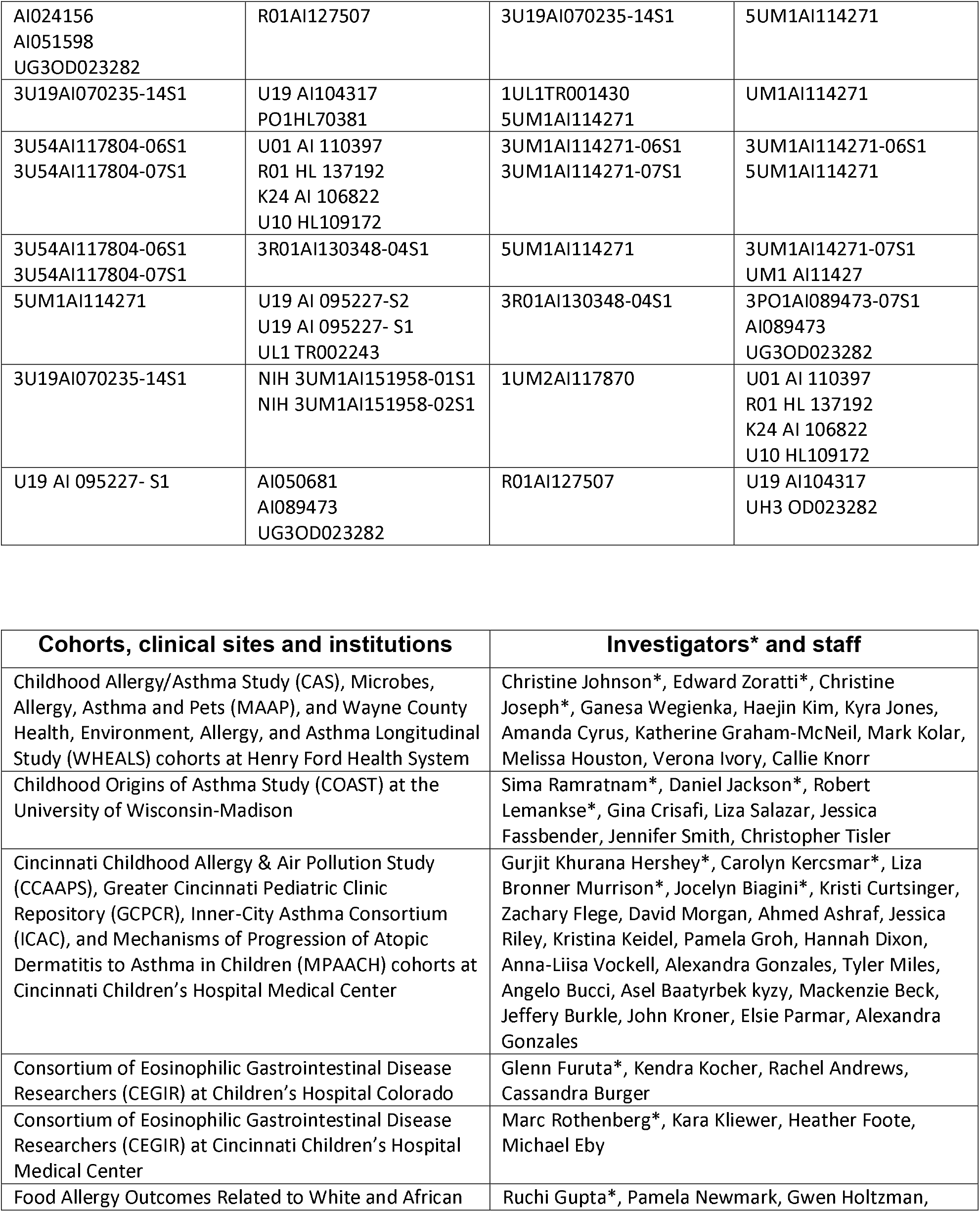
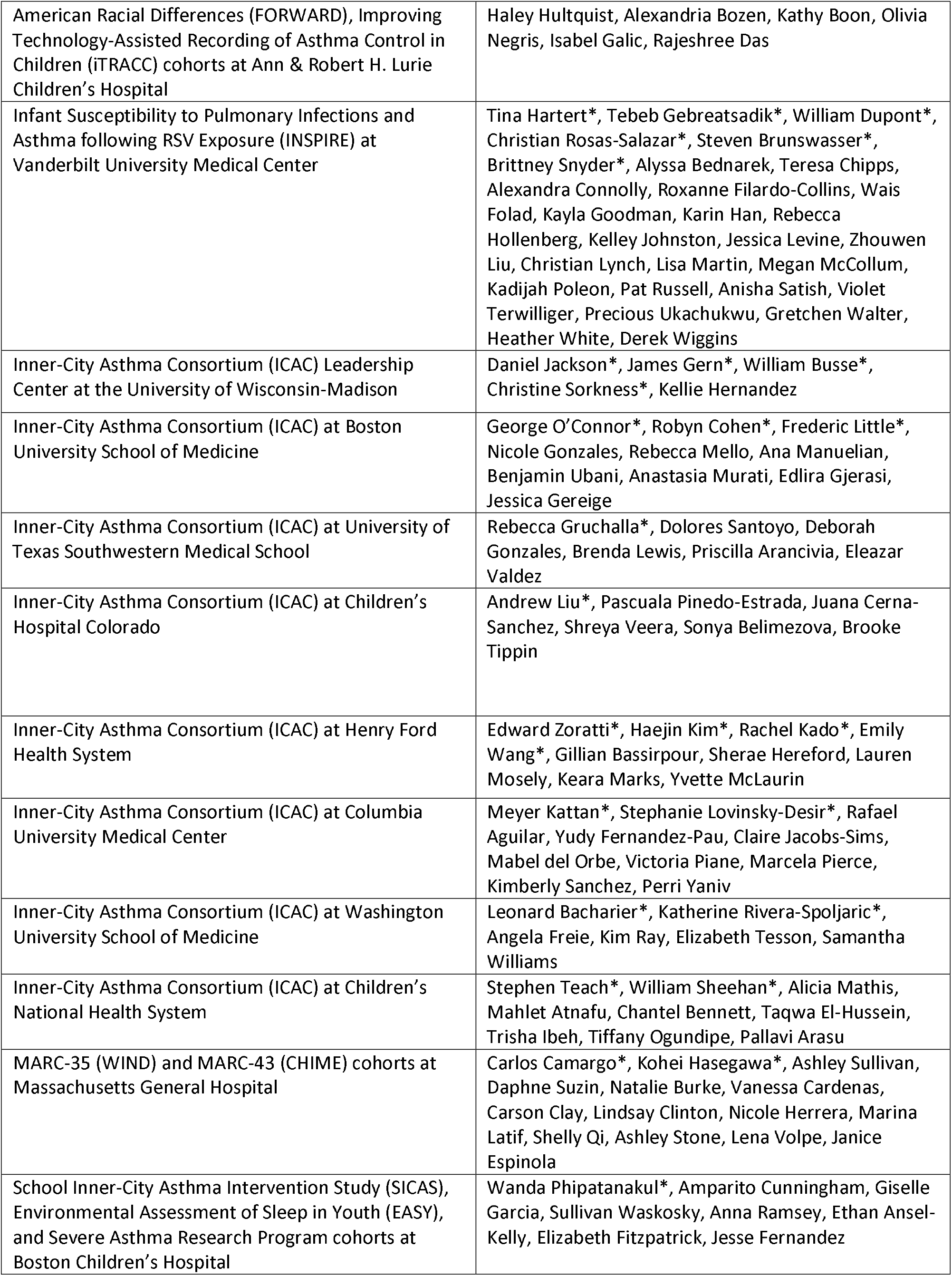
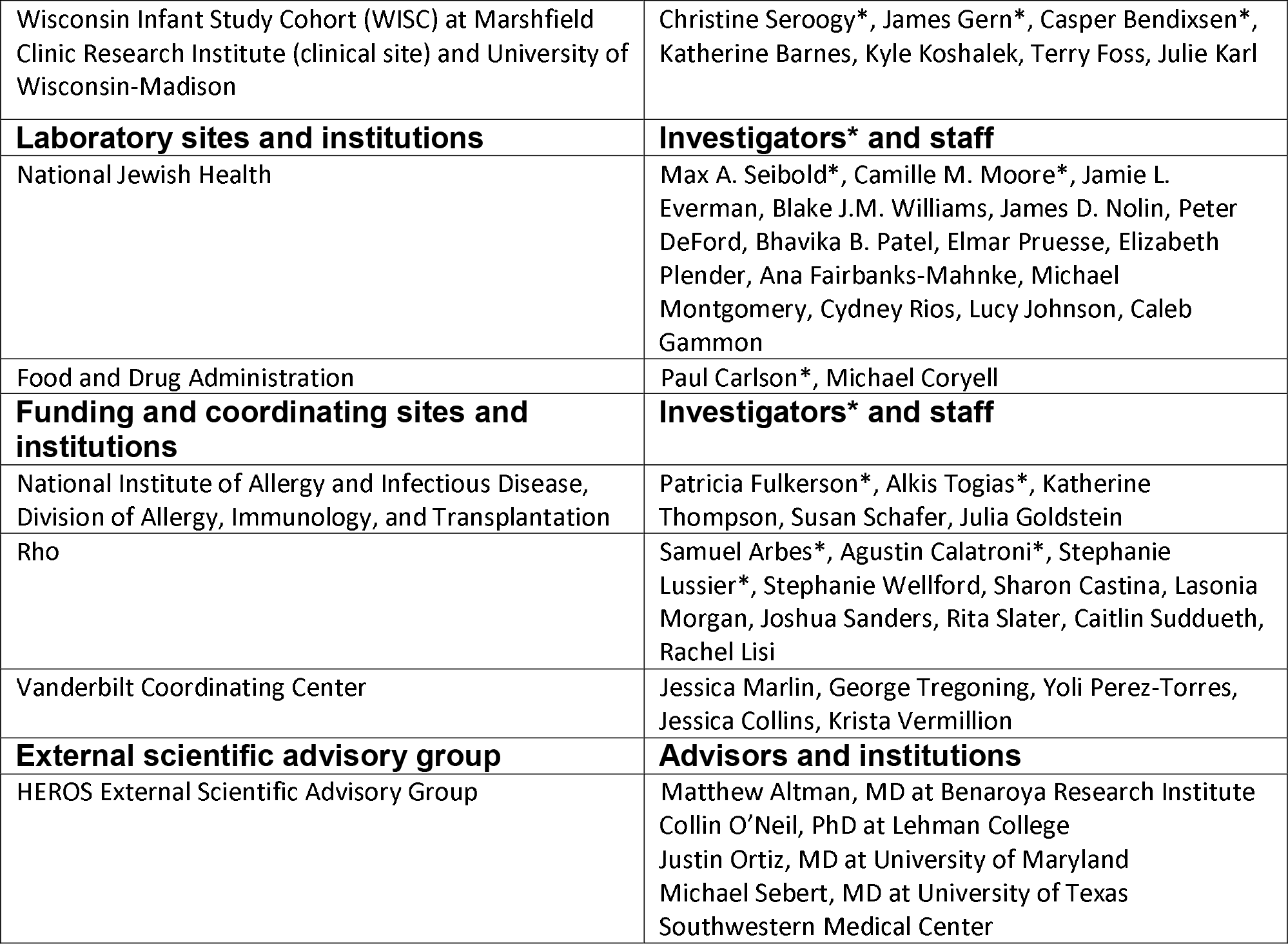

Also, special thanks to our colleagues who provided drafts of their developing Dutch COVID questionnaires for review and selection, and to the All of Us Data and Research Center’s Pilot Research team at Vanderbilt University Medical Center for their assistance in selecting survey items for inclusion, including Brandy Mapes and Rebecca L. Johnston, and to Barron Patterson, MD, our medical monitor.

