## Supplementary Materials for "Human Epidemiology and RespOnse to SARS-CoV-2 (HEROS): Objectives, Design and Enrollment Results of a 12-City Remote Observational Surveillance Study of Households with Children using Direct-to-Participant Methods"

**Contents**

**Supplemental methods**..……………………..…..………………...…………………………..…..2

**Supplemental tables**…………………………..…..………………....…………………………..…..4

**Supplemental figures**……………………………..………………...……………………….……..9

**Participant surveys link**………………………..………………...……………………………13

### Supplemental methods

**Study objectives**

The primary objective for HEROS was to determine the incidence of SARS-CoV-2 infection via detection of viral RNA in nasal secretions. Secondary objectives included: (1) to determine SARS-CoV-2 antibody development in children and their household members, (2) to compare SARS-CoV-2 infection status and virus-specific antibody development in children with asthma and other atopic conditions (e.g. eczema) versus children without asthma or other atopic conditions, (3) to determine the mucosal immune response to SARS-CoV-2 infection through gene expression profiling and to examine whether it is influenced by the presence of asthma and other atopic conditions, (4) to determine whether topical, systemic, or inhaled steroid use, as directed by the participants' health care provider, modifies the risk of SARS-CoV-2, the severity of infection, or the expression of the SARS-CoV-2 receptor, (5) to determine whether baseline demographic and environmental factors modify the risk of SARS-CoV-2 infection or the severity of infection, and (6) to assess whether a history of respiratory syncytial virus infection or bronchiolitis during infancy modifies severity of SARS-CoV-2 infection.

**Participants and enrollment**

An eligible household had to include a 21-years-old and younger individual who was or had been a participant in a NIH-funded asthma and other allergic diseases study (principal participant) and who was expected to remain in the United States with a caregiver for the duration of the study. In a few rare instances, individuals that had been participants in a NIH-funded allergic disease cohort and had aged into adulthood was enrolled in HEROS as a caregiver and their children were enrolled as the principal participant. Only individuals who had previously agreed to be contacted for other research were approached.

**Biospecimen collection and tracking**

All materials needed to collect and return a nasal swab, blood or stool collection for a household were provided in an individual kit boxes labeled on the outside by collection type. These boxes were customized depending on whether there were 2, 3 or 4 household members enrolled in the study. The initial collection box included a welcome letter for participants, instructions for sample collection, a color-coding chart for sample labeling and explanation of the kit contents. Supplies for the baseline nasal swab, blood and stool collections for the household, as well as nasal swab collection boxes for the next 3 collection timepoints were included. Also, a "Sick Kit" that included additional supplies for nasal and stool collections in the event a person in the family were to meet the criteria for an illness event requiring additional sampling. After getting some feedback from participants, the lead site study coordinator created an Unboxing Video where she showed the contents of each box and explained how to use them. A QR code linking to this video was included on the welcome letter and in the data entry system.

Three separate shipments were sent to households containing the nasal swab boxes as well as supplies for blood collection with the Tasso device and stool collection. A color-coding system was utilized to assign a specific color to each study participant in the household and color charts were provided to the households to assist with sample tracking. Each tube was labeled with a barcode and the color was typed on the label. Participants were instructed to take a picture of their samples with barcodes facing up. The barcodes were entered by the participant and the image of the samples were uploaded to the REDCap database. The images were used during the database clean up to reconcile barcode numbers entered by participants and samples that arrived at the biorepository. As the Tasso blood collection kits were validated in their shipping boxes, they were unable to be labeled directly on the tube by the supply vendor. The vendor provided a label in a zipped bag taped to the Tasso box. Participants packaged their samples in shipping materials provided and were instructed to mail the same or the next day via USPS.

Participants were instructed to swab the anterior nares for participants 10 years and younger and the interior turbinate for participants 11 years and older. If a study-specified symptom threshold was met (Supplemental Figure 1), an illness event was triggered for the household and additional illness-associated samples from all enrolled household participants were collected. At the time an illness event was triggered, an additional Tasso blood collection device would be shipped separately. If a household member not enrolled in HEROS developed symptoms consistent with COVID-19, the affected household member was provided the opportunity to review the consent-like study information sheet and enroll in the study to provide demographic, medical history, current medication, and the illness-associated samples.

**Data collection instruments**

The source instrumentation and corresponding items for HEROS baseline questionnaire and surveys were selected based on their relevance to the HEROS objectives and overall performance and/or extent of use when previously fielded. Minor modifications were made to items to ensure consistency in language or relevance to the study objectives. In addition, the baseline questionnaire and surveys contain some supplementary items that were developed by the study team based on review of COVID-19 relevant literature and consultation with study collaborators. The enrollment survey collected information on basic demographics and household features, and the enrollment and biweekly surveys included assessment of factors associated with exposure risks and risk-taking behaviors. A list of exposures was asked at the household level at enrollment and biweekly. The enrollment health assessment included: health history (allergies, allergies to food, hay fever/allergic rhinitis, asthma, autoimmune conditions, cancer, COPD, congestive heart failure, coronary artery disease, cystic fibrosis, emphysema, eczema (atopic dermatitis), heart attack, high cholesterol, hypertension, influenza, peripheral vascular disease, pneumonia, sleep apnea, type 1 or type 2 diabetes), current pregnancy, weight, height, quantified alcohol use, quantified smoking, detailed medication use, and home environment (number of household members, number of bedrooms, type of dwelling, pets). These included: Travel outside of the city or town that the family lives in, in-person work, in-person school, daycare, grocery store, in-person healthcare appointment, going out to eat, spending time with friends or family that do not live in the household, gatherings such as church or concerts, bringing home take-out food.

For those households that indicated a preference for communication via text or email, a link to the electronic surveys was sent to CG1 every week. A reminder was sent 24 and 48 hours later if the surveys were not completed. CG1 had 6 days to complete a survey before it was closed.

**Supplemental Table 1. Asthma and allergic disease focused cohorts included in HEROS, research cores, and associated funding**

| Cohort name | Area/region | Type of cohort or study site | Grant numbers |
|---|---|---|---|
| CAS | Southeastern MI, Midwestern US | Population-based cohort | AI024156<br>AI051598<br>UG3OD023282 |
| CCAAPS | Southwestern OH, Midwestern US | High risk cohort | 3U19AI070235-14S1 |
| CEGIR – Cincinnati | Southwestern OH, Midwestern US | Disease specific cohort - Eosinophilic gastrointestinal disorder | 3U54AI117804-06S1<br>3U54AI117804-07S1 |
| CEGIR – Denver | Central CO, Rocky Mountains US | Disease Specific cohort - Eosinophilic gastrointestinal disorder | 3U54AI117804-06S1<br>3U54AI117804-07S1 |
| CHIME | Northeastern MA, Northeastern US | Convenience cohort | R01AI127507 |
| COAST | Southwestern WI, Midwestern US | High risk cohort | U19 AI104317<br>PO1HL70381 |
| EASY | Northeastern MA, Northeastern US | Population-based and high risk cohort | U01 AI 110397<br>R01 HL 137192<br>K24 AI 106822<br>U10 HL109172 |
| FORWARD | Northeastern IL, Midwestern US | Disease specific cohort – Food allergy | 3R01AI130348-04S1 |
| GCPCR | Southwestern OH, Midwestern US | Convenience cohort | 3U19AI070235-14S1 |
| ICAC – Boston | Northeastern MA, Northeastern US | Convenience cohort; Disease specific cohort – Asthma; High-risk birth cohort – Children known to be at risk for developing asthma or allergic disease | 1UL1TR001430<br>5UM1AI114271 |
| ICAC – Cincinnati | Southwestern OH, Midwestern US | Convenience cohort; Disease specific cohort – Asthma | 3UM1AI114271-06S1<br>3UM1AI114271-07S1 |
| ICAC – Dallas | Central TX, Southwestern US | Convenience cohort; Disease specific cohort – Asthma | 5UM1AI114271 |
| ICAC – Denver | Central CO, Rocky Mountains US | Convenience cohort; Disease specific cohort – Asthma | 5UM1AI114271 |
| ICAC – Detroit | Southeastern MI, Midwestern US | Convenience cohort; Disease specific cohort – Asthma | UM1AI114271 |
| ICAC – New York | Metro NY, Northeastern US | Convenience cohort; Disease specific cohort – | 3UM1AI114271-06S1<br>5UM1AI114271 |

| | | | |
|---|---|---|---|
| | | Asthma; High-risk birth cohort – Children known to be at risk for developing asthma or allergic disease | |
| ICAC – St. Louis | Eastern MO, Midwestern US | Disease specific cohort – Asthma; High-risk birth cohort – Children known to be at risk for developing asthma or allergic disease | 3UM1AI14271-07S1<br>UM1 AI11427 |
| ICAC – Washington D.C. | Southwestern US | Convenience cohort; Disease specific cohort – Asthma | 5UM1AI114271 |
| INSPIRE | Middle TN, Southeastern US | Population-based birth cohort | U19 AI 095227-S2<br>U19 AI 095227- S1 |
| iTRACC | Northeastern IL, Midwestern US | Disease specific cohort – Asthma | 3R01AI130348-04S1 |
| MAAP | Southeastern MI, Midwestern US | Population-based birth cohort | 3PO1AI089473-07S1<br>AI089473<br>UG3OD023282 |
| MPAACH | Southwestern OH, Midwestern US | Disease specific cohort – Atopic dermatitis | 3U19AI070235-14S1 |
| National Jewish Health | Central CO, Rocky Mountains US | N/A | NIH 3UM1AI151958-01S1<br>NIH 3UM1AI151958-02S1 |
| Rho | Eastern NC, Southeastern US | N/A | 1UM2AI117870 |
| SARP | Northeastern MA, Northeastern US | Disease specific cohort – Asthma | U01 AI 110397<br>R01 HL 137192<br>K24 AI 106822<br>U10 HL109172 |
| SICAS | Northeastern MA, Northeastern US | Disease specific cohort – Asthma | U01 AI 110397<br>R01 HL 137192<br>K24 AI 106822<br>U10 HL109172 |
| Vanderbilt Coordinating Center | Middle TN, Southeastern US | Data coordinating center | U19 AI 095227- S1 |
| WHEALS | Southeastern MI, Midwestern US | Population-based birth cohort | AI050681<br>AI089473<br>UG3OD023282 |
| WIND | Northeastern MA, Northeastern US | Disease specific cohort - severe (hospitalized) bronchiolitis during infancy | R01AI127507 |
| WISC | North Central WI, Midwestern US | Children enrolled from farm families and non-farm families | U19 AI104317<br>UH3 OD023282 |

**Supplemental Table 2. Self-reported household clinical and social characteristics at enrollment**

| | Completed, N = 3,081[1] | Withdrawn, N = 2,517[1] | Overall, N = 5,598[1] | p-value[2] |
|---|---|---|---|---|
| **Doctor Indicated COVID** | 37 (1.2%) | 37 (1.5%) | 74 (1.3%) | 0.4 |
| **Allergies (Hay Fever, Allergic Rhinitis)** | 1,383 (45%) | 1,123 (45%) | 2,506 (45%) | 0.8 |
| **Allergies to Food** | 489 (16%) | 395 (16%) | 884 (16%) | 0.9 |
| **Asthma** | 830 (27%) | 843 (33%) | 1,673 (30%) | <0.001 |
| **Eczema** | 555 (18%) | 440 (17%) | 995 (18%) | 0.6 |
| **No Reported Allergic Disease** | 1,248 (41%) | 965 (38%) | 2,213 (40%) | 0.10 |
| **Alcohol Consumption Frequency** | | | | <0.001 |
| Four or more times a week | 138 (9.3%) | 82 (7.0%) | 220 (8.3%) | |
| Monthly or less | 433 (29%) | 406 (35%) | 839 (32%) | |
| Never | 318 (21%) | 311 (27%) | 629 (24%) | |
| Prefer not to answer | 20 (1.3%) | 26 (2.2%) | 46 (1.7%) | |
| Two to four times a month | 296 (20%) | 226 (19%) | 522 (20%) | |
| Two to three times a week | 283 (19%) | 120 (10%) | 403 (15%) | |
| N Missing | 1,593 | 1,346 | 2,939 | |
| **Smoking Cigarette/Cigar Frequency** | | | | <0.001 |
| Daily | 67 (3.8%) | 145 (10%) | 212 (6.7%) | |
| Less than Daily | 55 (3.1%) | 61 (4.4%) | 116 (3.7%) | |
| Not at all | 1,627 (93%) | 1,175 (84%) | 2,802 (89%) | |
| Prefer not to answer | 8 (0.5%) | 13 (0.9%) | 21 (0.7%) | |
| N Missing | 1,324 | 1,123 | 2,447 | |
| **Electronic Nicotine Product Frequency** | | | | 0.058 |
| Daily | 17 (1.0%) | 21 (1.5%) | 38 (1.2%) | |
| Less than Daily | 15 (0.9%) | 25 (1.8%) | 40 (1.3%) | |
| Not at all | 1,717 (98%) | 1,340 (96%) | 3,057 (97%) | |
| Prefer not to answer | 8 (0.5%) | 6 (0.4%) | 14 (0.4%) | |
| N Missing | 1,324 | 1,125 | 2,449 | |

[1]n (%)

[2]Pearson's Chi-squared test

| **Supplemental Table 3. Biospecimen collection adherence by caregiver demographics** | | |
|---|---|---|
| | **% Blood Samples mean (sd)** | **% Nasal Samples median (q1, q3)** |
| **Sex** | | |
| Female | 54% (41%) | 57% (10%, 86%) |
| Intersex | 100% (NA) | 64% (64%, 64%) |
| Male | 66% (40%) | 79% (43%, 93%) |
| Prefer Not to Answer | 100% (NA) | 64% (64%, 64%) |
| Unknown | 0% (NA) | 0% (0%, 0%) |
| **Race** | | |
| American Indian or Alaska Native | 49% (44%) | 43% (0%, 75%) |
| Asian | 70% (35%) | 82% (57%, 93%) |
| Black or African American | 37% (40%) | 20% (0%, 64%) |
| More than One Race | 48% (44%) | 43% (10%, 79%) |
| Native Hawaiian or Other Pacific Islander | 50% (71%) | 55% (32%, 78%) |
| Prefer Not to Answer | 51% (42%) | 48% (0%, 80%) |
| Unknown | 0% (NA) | 0% (0%, 0%) |
| White or Caucasian | 65% (38%) | 76% (30%, 93%) |
| **Ethnicity** | | |
| Hispanic | 54% (42%) | 50% (10%, 79%) |
| Non-Hispanic | 55% (41%) | 62% (10%, 86%) |
| Prefer Not to Answer | 41% (41%) | 15% (0%, 80%) |
| Unknown | 0% (NA) | 0% (0%, 0%) |
| **Smoking History** | | |
| Daily | 33% (37%) | 10% (0%, 64%) |
| Less than Daily | 43% (44%) | 40% (0%, 79%) |
| Not at all | 57% (41%) | 64% (10%, 86%) |
| Prefer not to answer | 44% (40%) | 29% (0%, 52%) |

| Supplemental Table 4. Adverse events | |
|---|---|
| | N = 5,598[1] |
| **Toxicity Grade** | |
| No AE Reported | 5,579 (99.66%) |
| Grade 1 (Mild) | 15 (0.27%) |
| Grade 2 (Moderate) | 1 (0.02%) |
| Grade 3 (Severe and undesirable) | 3 (0.05%) |
| **Relation to Blood Collection** | |
| Not Related | 1 (5.26%) |
| Possibly Related | 5 (26.32%) |
| Definitely Related | 13 (68.42%) |
| **Relation to Nasal Swab Collection** | |
| Not Related | 17 (89.47%) |
| Possibly Related | 1 (5.26%) |
| Definitely Related | 1 (5.26%) |
| **Concussion** | 1 (0.02%) |
| **Contusion** | 1 (0.02%) |
| **Dermatitis contact** | 1 (0.02%) |
| **Dizziness** | 2 (0.04%) |
| **Epistaxis** | 2 (0.04%) |
| **Facial bones fracture** | 1 (0.02%) |
| **Syncope** | 6 (0.11%) |
| **Vascular catheter specimen collection** | 1 (0.02%) |
| **Vessel puncture site bruise** | 3 (0.05%) |
| **Vessel puncture site hemorrhage** | 1 (0.02%) |
| **Vessel puncture site hypoaesthesia** | 1 (0.02%) |
| **Vessel puncture site pain** | 2 (0.04%) |
| **Vessel puncture site rash** | 1 (0.02%) |
| **Vessel puncture site swelling** | 2 (0.04%) |

[1]n (%)

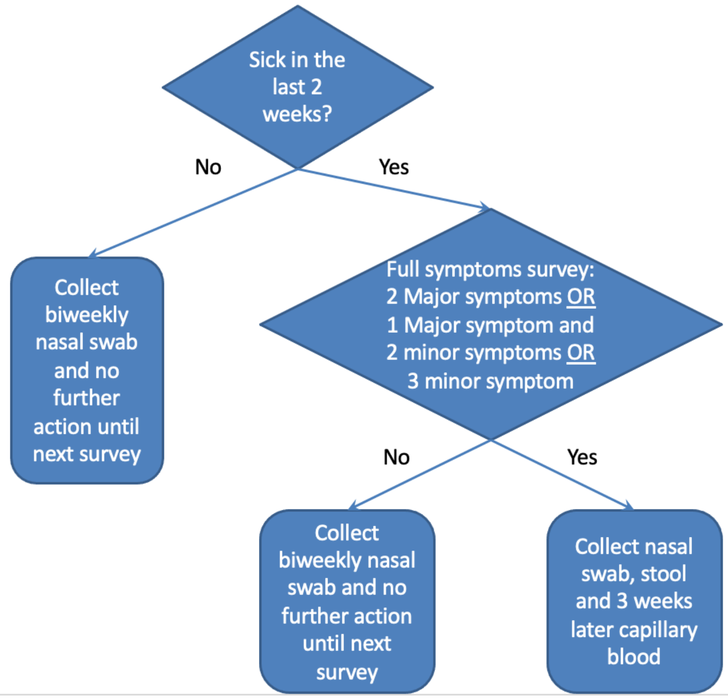


Algorithm: (Were you sick within the last 2 weeks? = YES) AND (2 Major symptoms OR 1 Major symptom and 2 minor symptoms OR 3 minor symptoms)

**MAJOR SYMPTOMS**
Did you have a fever or felt feverish (chills, sweating) in the last 2 weeks? =YES
Did you have a cough in the last 2 weeks? = YES
Did you have any shortness of breath at any time during the last 2 weeks= YES
Did you have any pain or discomfort in your chest within the last 2 weeks?  = YES
Did you feel tired or fatigued during the last 2 weeks? = YES
Did you experience any diarrhea within the last 2 weeks? = YES

**MINOR SYMPTOMS**
Did you have any body aches during the last 2 weeks? = YES
Did you have any cold or flu-like symptoms (such as a sore throat, runny nose, or congestion)=YES
Did you have any headaches at any time during the last 2 weeks=YES
Did you have a problem with your ability to smell, such as not being able to smell things or things not smelling the way they are supposed to within the last 2 weeks? = YES
Did you have a problem with your ability to taste sweet, sour, salty, or bitter foods and drinks within the last 2 weeks? = YES
Did you experience any nausea (feeling you might vomit) within the last 2 weeks? = YES
Did you experience any belly pain at any time within the last 2 weeks? = YES
Did you have red or pink eyes at any time during the last 2 weeks = YES
For age < 2 years, irritability
For age < 2 years, difficulty feeding
For age < 2 years, altered sleep (more or less)
For age < 2 years, pulling at ears
For age < 6 years, Did you have any ear pain at any time during the last 2 weeks

**Supplemental Figure 1. Symptom algorithm to trigger an illness event and additional household biospecimen collection based on survey question responses.**

**Supplemental Figure 2. Flow diagram of remote data and biospecimen management.**

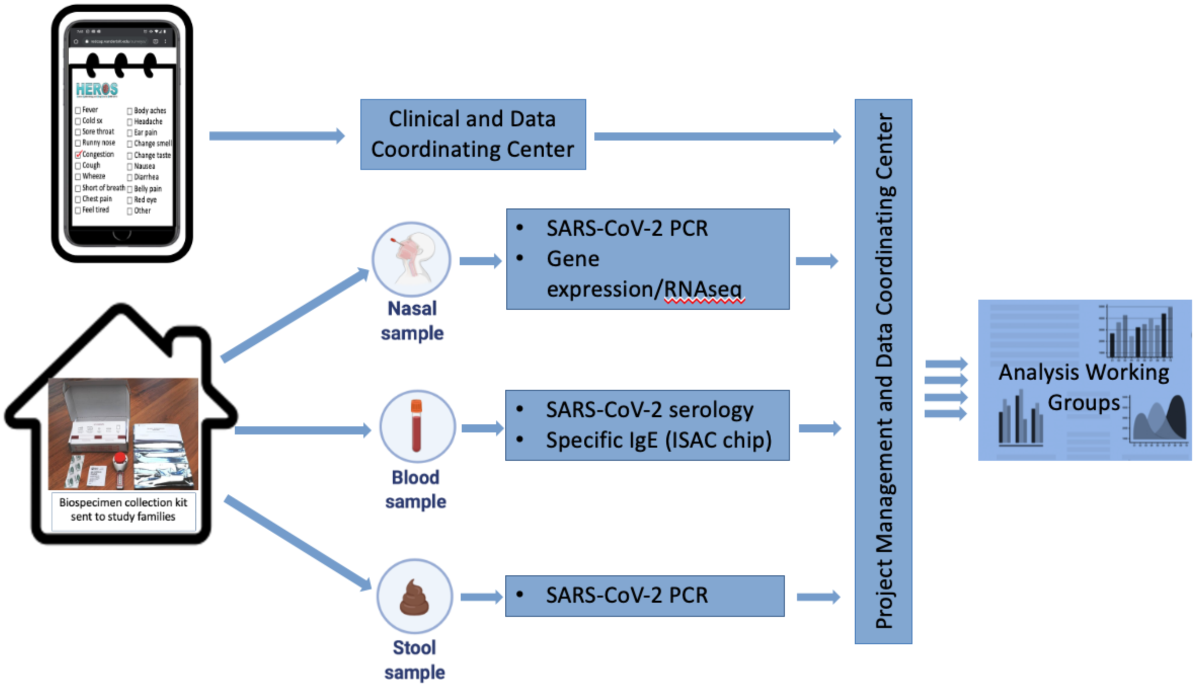

**Supplemental Figure 3. Household collection of biweekly nasal samples over the study period.**

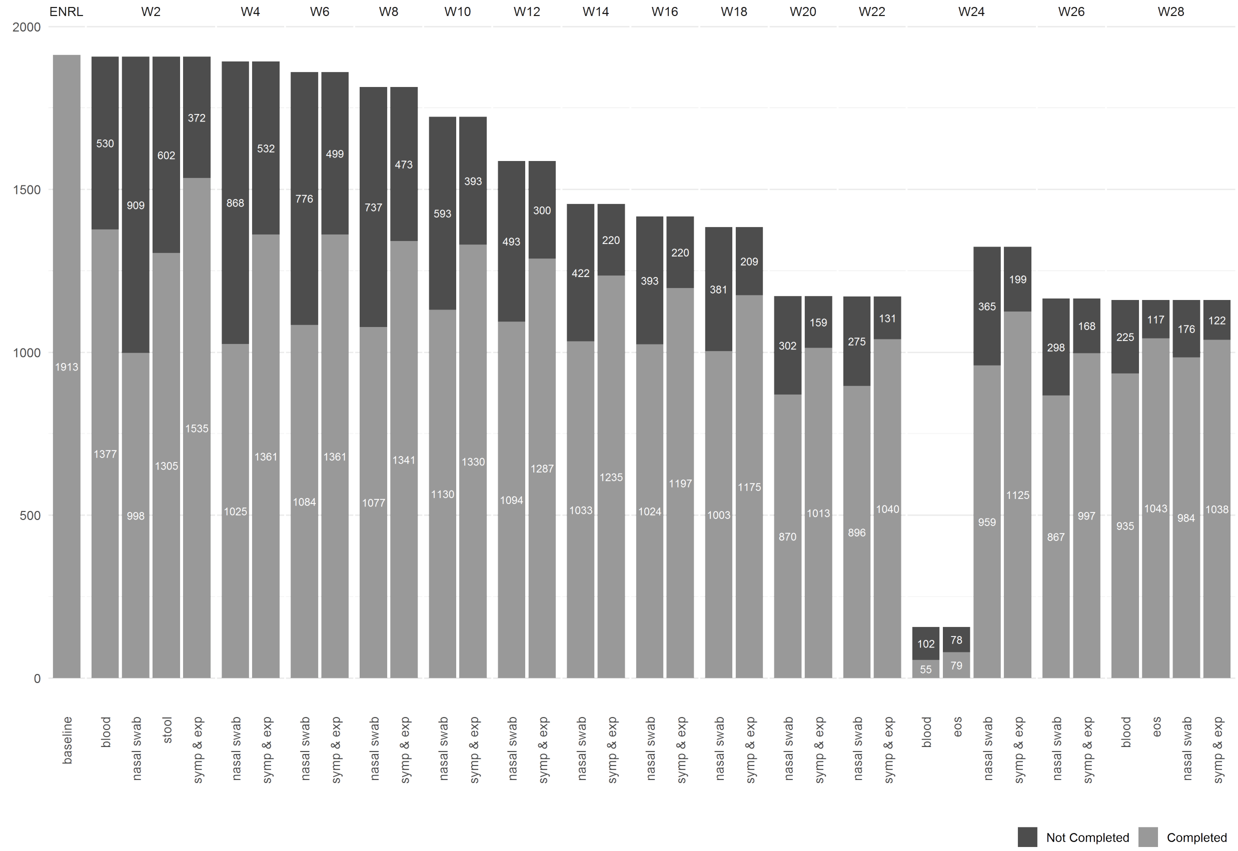


Baseline – enrollment survey
Blood – Capillary blood collection
Nasal swab – biweekly nasal collection
EOS – end of study survey
Symp & exp – biweekly survey of symptoms, behaviors and exposures

**Supplemental Figure 4. Cumulative household withdrawal over study period relative to enrollment.**

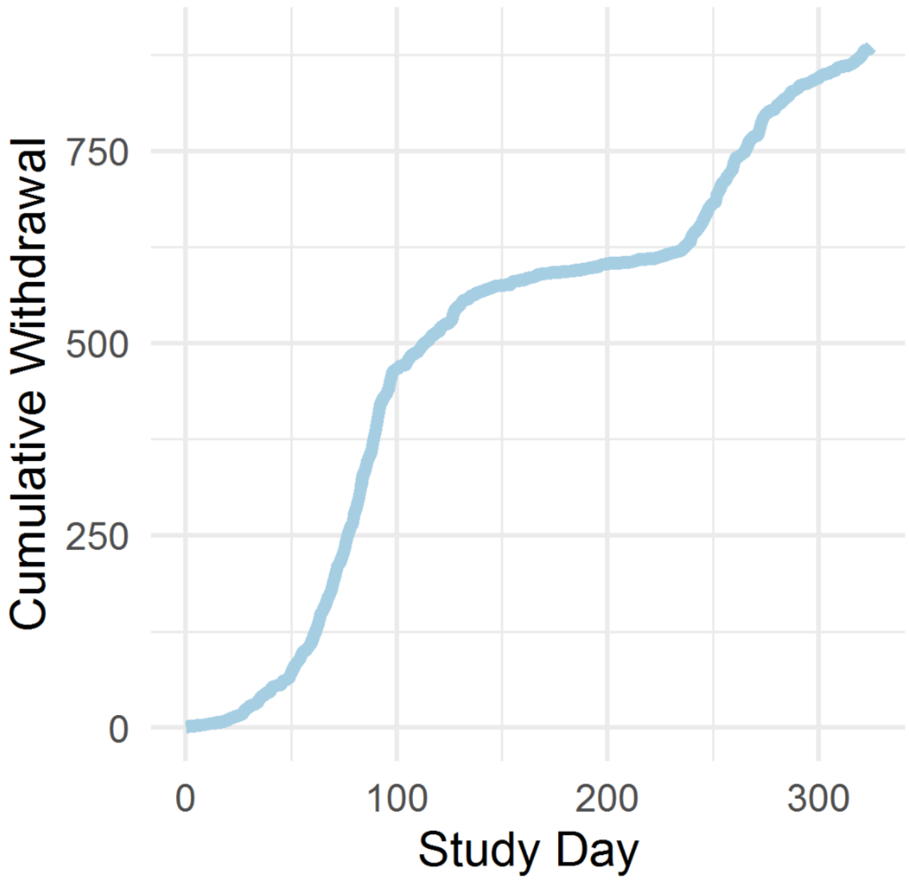

**Participant Surveys Link:** https://www.vumc.org/heros/survey-instruments

**Downloads:**

HEROS Registration Questionnaire (English)
HEROS Baseline Enrollment Questionnaire (English)
HEROS Weekly Health Check (English)
HEROS Every-Other-Week Questionnaire (English)
HEROS Child Specific Bi-Weekly Questions (English)
HEROS Sample Collection (English)
End of study survey (English)

HEROS Registration Questionnaire (Spanish)
HEROS Baseline Enrollment Questionnaire (Spanish)
HEROS Weekly Health Check Questionnaire (Spanish)
HEROS Every-Other-Week Questionnaire (Spanish)
HEROS Child Specific Bi-Weekly Questionnaire (Spanish)
HEROS Sample Collection (Spanish)
End of study survey (Spanish)
